# Predicting the 10-year risk of cardiomyopathy in long-term survivors of childhood cancer: a report from the St. Jude Lifetime Cohort and the Childhood Cancer Survivor Study

**DOI:** 10.1101/2024.10.24.24316064

**Authors:** Kateryna Petrykey, Yan Chen, Achal Neupane, Jennifer French, Huiqi Wang, Haoxue Xiang, Stephanie B. Dixon, Chris Vukadinovich, Cindy Im, Matthew J. Ehrhardt, Daniel A. Mulrooney, Noha Sharafeldin, Xuexia Wang, Rebecca M. Howell, John L. Jefferies, Paul W. Burridge, Kevin C. Oeffinger, M. Monica Gramatges, Smita Bhatia, Leslie L. Robison, Kirsten K. Ness, Melissa M. Hudson, Eric J. Chow, Gregory T. Armstrong, Yutaka Yasui, Yadav Sapkota

## Abstract

**Purpose:** Considering the heightened risk of cancer treatment-related cardiomyopathy and cardiac death in long-term survivors of childhood cancer, we aimed to develop and validate a clinically-applicable risk prediction model for cardiomyopathy.

**Patients and Methods:** Childhood cancer survivors from St. Jude Lifetime Cohort, (SJLIFE, model-development; n=3,479; median age 32.3 years, IQR 24.4-40.9) and Childhood Cancer Survivor Study (CCSS, model-validation; n=6,875; median age 33.2 years, IQR 27.9-38.9) were assessed for demographic and cardiovascular risk factors, treatment exposures, and polygenic risk scores (PRSs) for cardiomyopathy, heart failure, cardiac structure and function, and anthracycline-related cardiomyopathy risk. Multivariable Poisson regression predicted the 10-year risk of cardiomyopathy (CTCAE grade ≥3: requiring heart failure medications or heart transplantation or leading to death) following baseline visit/survey. Model performance was assessed by area under the receiver operating characteristic curve (AUC).

**Results:** Cardiomyopathy was clinically identified in 75 (2.2%, SJLIFE) and self-reported in 87 (1.3%, CCSS) survivors within 10 years of the baseline assessment. AUC of a clinical model with sex, age at cancer diagnosis, cumulative anthracycline and mean heart radiation doses was 0.833 (SJLIFE) and 0.812 (CCSS). Age at baseline, hypertension and genetic ancestry showed associations with higher cardiomyopathy rates in SJLIFE but did not increase AUC in CCSS (0.812). Adding PRSs for hypertrophic cardiomyopathy and left ventricular end-systolic volume improved AUC in CCSS (0.822; *P*=0.016). Compared to existing survivorship-care guidelines, the PRS model classified fewer survivors as high-risk or moderate-risk, while identifying survivors in those categories as having 1.5-times greater risk.

**Conclusion:** We developed and validated a model with highest-to-date performance for estimating the 10-year risk of cardiomyopathy in survivors of childhood cancer. Results could enhance identification of at-risk survivors beyond current guidelines.

## INTRODUCTION

In the United States, over 85% of children diagnosed with cancer survive for at least five years, resulting in nearly half a million childhood cancer survivors today^1^. However, these survivors often face chronic health issues from their cancer and treatments, increasing their risk of early death^2,3^. Cardiovascular disease, including cardiomyopathy and heart failure, is the leading non-cancer cause of mortality in this population^4–6^, with prevalence increasing with age^7^. Previous exposure to cardiotoxic treatments, such as anthracycline chemotherapy and chest-directed radiotherapy, are known risk factors. Additionally, cardiovascular risk factors (CVRFs), such as hypertension, dyslipidemia, and diabetes, further exacerbate the cardiac risks associated with these cancer treatments^8,9^.

Risk-prediction models guide primary cardiovascular prevention and treatment in the general population^10–13^. However, these models likely underestimate risks in childhood cancer survivors because they do not incorporate cardiotoxic cancer therapy exposures. Two previous risk prediction models for heart failure (HF) in survivors incorporated cancer treatment and/or CVRFs but were based on self-reported data from the CCSS and did not consider age or race as predictors^14,15^. Compared to clinically-assessed outcomes, self-reported data can lead to misclassification and imprecise predictor estimates, adversely impacting model performance. In the general population, genetic variants are associated with cardiomyopathy^16^, HF^17^, and cardiac structure and function^16,18^. In survivors, genetic variants have been associated with anthracycline-related cardiomyopathy^19–24^. However, the role of these genetic factors in predicting cardiomyopathy risk among survivors has not been fully explored.

This study aimed to utilize unique data from the clinically-assessed SJLIFE cohort of childhood cancer survivors to develop accurate and clinically-applicable risk prediction models for cardiomyopathy. These models incorporated survivors’ characteristics including age at baseline visit, treatment exposures, and inherited genetic variation. Models were independently validated using survivors from the CCSS.

## PATIENTS AND METHODS

### Study population

Participants were from two large cohort studies of childhood cancer survivors in North America, SJLIFE^25,26^ and CCSS^27,28^. SJLIFE is a retrospectively constructed cohort initiated in 2007, with prospective clinical assessments of ≥5-year survivors of childhood cancer treated at St. Jude Children’s Research Hospital (SJCRH) since 1962. CCSS is also a retrospectively constructed cohort study with prospective follow-up of ≥5-year survivors diagnosed before age 21 and treated for various cancers at 31 participating institutions in the U.S and Canada between 1970 and 1999. Here, we included survivors in SJLIFE (model-development) and CCSS (model-validation; independent of those also in SJLIFE) who had genome-wide genetic data and completed at least one campus visit or questionnaire. Institutional review boards at SJCRH and each of the CCSS participating centers approved the study, and all participants provided written informed consent.

### Definitions of outcomes

Cardiomyopathy in SJLIFE was clinically assessed based on left ventricular ejection fraction (LVEF) and pharmacologic treatment and graded per a modified version of the National Cancer Institute’s Common Terminology Criteria for Adverse Events (CTCAE v4.03)^29^. Participants in CCSS completed a multi-item questionnaire at baseline and follow-up evaluations that included age at onset of HF based on self-report of medical diagnosis and pharmacological treatment. Questionnaire items related to HF in CCSS were classified and graded using the CTCAE methodology^8,30^. Survivors in both SJLIFE and CCSS with CTCAE grade ≥3 were considered affected. Detailed CTCAE grading criteria for both cohorts are provided in Supplementary Table 1.

### Cancer treatment exposures and CVRFs

Information pertaining to chemotherapy and radiotherapy exposures was abstracted from medical records (Supplementary Methods). The cumulative anthracycline dose (in mg/m^2^) was determined by doxorubicin toxicity equivalence^31^, and mean radiation dose to the heart for each survivor was calculated using established radiation dosimetry methodologies^32–34^. CVRFs including hypertension, dyslipidemia, and diabetes were clinically-assessed in SJLIFE (comprising laboratory measurements and blood pressure values in addition to pharmacologic treatment) and were self-reported in CCSS. These CVRFs were defined as CTCAE grade ≥2 in both cohorts.

### Polygenic risk scores

Based on studies in the general population and childhood cancer survivors, we considered multiple PRSs for cardiomyopathies (dilated [DCM] and hypertrophic [HCM]), HF, measures of cardiac structure and function, and anthracycline-related cardiomyopathy. Specifically, we considered six separate PRSs for DCM (PRS_DCM_)^16^, HCM (PRS_HCM_)^16^, HF (PRS_HF_)^17^, LVEF (PRS_LVEF_)^18^, LV end-systolic volume index (PRS_LVESVi_)^18^ and anthracycline-related cardiomyopathy (PRS_ACT_)^19–24^. Details regarding genotyping, genetic ancestry determination, and the PRS calculation are described in Supplementary Methods.

### Statistical Analysis

Multivariable Poisson regression models were fit sequentially to the SJLIFE model-development cohort to evaluate associations with incident cardiomyopathy, starting with variables selected *a priori* from the existing HF risk prediction models previously developed in the CCSS^35,36^. The variables included in this clinical model were age at primary cancer diagnosis, sex, cumulative anthracycline dose, and mean heart radiation dose. Specifically, we predicted the incidence of CTCAE grade ≥3 cardiomyopathy over a 10-year period, incorporating an offset to account for variations in follow-up length: *i.e.*, the offset was the time between a survivor’s baseline assessment (or cohort entry) and the earliest of his/her last follow-up, 10 years after the baseline, or death. This approach allowed us to scale each survivor’s risk according to the actual follow-up duration, ensuring accurate rate estimation while adjusting for differences in follow-up length. Survivors with grade ≥3 cardiomyopathy (or grade ≥3 HF) prior to baseline, or those whose first cardiomyopathy was grade 2 and occurred before baseline, were excluded from the analysis. The subsequent models included age and prevalent CVRFs at baseline. The CVRFs retained in the model were selected through a backward selection approach. Genetic ancestry was then entered into the model (with a sensitivity analysis that replaced genetic ancestry with self-reported race), followed by an addition of each of the six PRSs as z-scores – PRS_DCM_, PRS_HCM_, PRS_LVEF_, PRS_LVESVi_, PRS_HF_, and PRS_ACT_. The PRSs with *P*<0.1 in this univariate analysis were added to the model simultaneously.

Receiver operating characteristic (ROC) analyses were performed to assess the predictive ability, measured using the area under the ROC curves (AUC), of the following five models – model 1 (clinical model); model 2 (model 1 with age at baseline); model 3 (model 2 with CVRFs at baseline); model 4 (model 3 with genetic ancestry or self-reported race); and model 5 (model 4 with the PRSs). The prediction models 1-5 developed based on SJLIFE participants were validated using the independent model-validation cohort of CCSS participants, calculating AUCs of the exact same models developed from SJLIFE using the same predictors and the model parameter values fixed as the estimates from SJLIFE. The statistical significance of AUC improvement by including additional predictor(s) over the model without them was assessed using the DeLong’s test^37^. All five models, including the univariate analyses of PRSs, were also assessed among survivors of European ancestry in both cohorts, since over 80% of survivors in SJLIFE/CCSS were of European ancestry, and the PRSs were primarily derived from studies involving individuals of European ancestry.

The CCSS model-validation cohort used for all model validations was required to have genome-wide genotype data, which is available for only about one-third of the entire CCSS cohort. For generalizability, however, models 1-4 without requiring the genotype-data were also evaluated in the entire CCSS cohort (n=21,352; 217 with CTCAE grade ≥3 HF after baseline). Additionally, the models were evaluated among survivors exposed to cardiotoxic therapies (anthracyclines and/or chest-directed radiotherapy [chest RT]), and those in the following risk groups based on the current International Late Effects of Childhood Cancer Guideline Harmonization Group (IGHG)^38^ recommendations: high-risk group (≥250 mg/m^2^ anthracyclines or ≥30 Gray chest RT, or a combination of ≥100 mg/m^2^ anthracyclines and ≥15 Gray chest RT); moderate-risk group (100 mg/m^2^≤anthracyclines<250 mg/m^2^ or 15 Gray≤chest RT<30 Gray); and low-risk group (0 mg/m^2^<anthracyclines<100 mg/m^2^ or 0 Gray<chest RT<15 Gray).

To assess the calibration of the best-performing model, the predicted probability of developing cardiomyopathy in the 10-year period from the baseline for each survivor (assuming no death) was compared to the observed cumulative incidence of cardiomyopathy. Based on these predicted probabilities, survivors were then categorized into “high” (predicted probability>10%), “moderate” (5%≤predicted probability≤10%), and “low” (predicted probability<5%) risk groups. This <5% threshold is the criterion used to identify low-risk individuals for atherosclerotic cardiovascular disease in the general population, as defined by the pooled cohort equations^39^.The 10-year cumulative incidence of cardiomyopathy in each risk group was estimated and compared to those of the IGHG-based risk groups.

## RESULTS

A total of 3,479 survivors in SJLIFE were available for model development and 6,875 survivors in CCSS were included for model validation. Over the 10 years following baseline, cardiomyopathy was clinically identified in 75 (2.2%) SJLIFE survivors and self-reported in 87 (1.3%) CCSS survivors. Clinical, demographic, and treatment characteristics of study participants are provided in Table 1.

**Table 1.** Demographic and clinical characteristics of childhood cancer survivors from the St. Jude Lifetime Cohort and Childhood Cancer Survivor Study

| Characteristics | SJLIFE discovery cohort |  | CCSS validation cohort |  |
| --- | --- | --- | --- | --- |
|  | Cardiomyopathy within 10 years from baseline (n=75) | Without cardiomyopathy (n=3,404) | Cardiomyopathy within 10 years from baseline (n=87) | Without cardiomyopathy (n=6,788) |
| Age at the primary diagnosis (years) |  |  |  |  |
| ≤5 | 22 (29.3) | 1,523 (44.7) | 8 (9.2) | 2,561 (37.7) |
| >5-10 | 15 (20.0) | 717 (21.1) | 18 (20.7) | 1,545 (22.8) |
| >10-15 | 20 (26.7) | 688 (20.2) | 21 (24.1) | 1,517 (22.3) |
| >15-21 | 18 (24.0) | 476 (14.0) | 40 (46.0) | 1,165 (17.2) |
| Sex |  |  |  |  |
| Male | 45 (60.0) | 1,719 (50.5) | 31 (35.6) | 3,252 (47.9) |
| Female | 30 (40.0) | 1,685 (49.5) | 56 (64.4) | 3,536 (52.1) |
| Cumulative anthracycline dose (mg/m<sup>2</sup>)* |  |  |  |  |
| None | 24 (32.0) | 1,540 (45.2) | 34 (39.1) | 4,022 (59.3) |
| >0-100 | 7 (9.3) | 741 (21.8) | 2 (2.3) | 443 (6.5) |
| >100-250 | 20 (26.7) | 807 (23.7) | 8 (9.2) | 1,236 (18.2) |
| >250 | 24 (32.0) | 316 (9.3) | 43 (49.4) | 1,087 (16.0) |
| Mean heart radiation exposure dose (Gray) |  |  |  |  |
| None | 23 (30.7) | 1,882 (55.3) | 21 (24.1) | 3,126 (46.1) |
| ≤5 | 22 (29.3) | 1,016 (29.8) | 25 (28.7) | 2,218 (32.7) |
| >5-15 | 6 (8.0) | 214 (6.3) | 5 (5.7) | 588 (8.7) |
| >15-35 | 18 (24.0) | 282 (8.3) | 31 (35.6) | 811 (11.9) |
| >35 | 6 (8.0) | 10 (0.3) | 5 (5.7) | 45 (0.7) |
| Age at baseline (years) |  |  |  |  |
| ≤25 | 10 (13.3) | 1477 (43.4) | 22 (25.3) | 3480 (51.3) |
| >25-35 | 25 (33.3) | 1,195 (35.1) | 34 (39.1) | 2,633 (38.8) |
| >35-45 | 28 (37.4) | 561 (16.5) | 31 (35.6) | 662 (9.8) |
| >45 | 12 (16.0) | 171 (5.0) | 0 (0.0) | 13 (0.2) |
| Hypertension** |  |  |  |  |
| Yes | 25 (33.3) | 464 (13.6) | 10 (11.5) | 283 (4.2) |
| No | 50 (66.7) | 2,940 (86.4) | 77 (88.5) | 6,505 (95.8) |
| Dyslipidemia** |  |  |  |  |
| Yes | 21 (28.0) | 412 (12.1) | 3 (3.4) | 157 (2.3) |
| No | 54 (72.0) | 2,992 (87.9) | 84 (96.6) | 6,631 (97.7) |
| Diabetes** |  |  |  |  |
| Yes | 9 (12.0) | 231 (6.8) | 1 (1.1) | 107 (1.6) |
| No | 66 (88.0) | 3,173 (93.2) | 86 (98.9) | 6,681 (98.4) |
| Genetic ancestry |  |  |  |  |
| European | 60 (80.0) | 2,709 (79.6) | 81 (93.1) | 6,147 (90.6) |
| African | 14 (18.7) | 512 (15.0) | 3 (3.4) | 153 (2.3) |
| Other | 1 (1.3) | 183 (5.4) | 3 (3.4) | 488 (7.2) |

\*Doxorubicin toxicity equivalence
\*\*At baseline

The fit of the multivariable Poisson-regression models 1-4 based on SJLIFE survivors is shown in Supplementary Table 3. Model 2 revealed a significant association between the 10-year cardiomyopathy incidence and baseline age >35-45 years (relative rate [RR]=2.40; *P*=0.027) and ≥45 years (RR=3.35; *P*=0.014) compared to age ≤25 years. In model 3, the only CVRF retained was hypertension (RR=2.20; *P*=2.7×10^-3^). In model 4, African genetic ancestry was suggestively associated (RR=1.70; *P*=0.078), with similar results for self-reported Black race (RR=1.66; *P*=0.084). Adding each of the six PRSs to model 4 showed none with *P*<0.1 for all survivors (Supplementary Table 4). However, in survivors of European ancestry, two PRSs showed *P*<0.1 (PRS_LVESVi_: RR per standard deviation=1.26; *P*=0.082 and PRS_HCM_: RR per standard deviation=0.72; *P*=0.024). Considering over 80% of survivors in both SJLIFE and CCSS cohorts were of European ancestry, these two PRSs were added simultaneously to model 5 (Table 2).

**Table 2.** The final multivariable Poisson regression model estimating the cardiomyopathy incidence rate over the next 10 years from baseline among childhood cancer survivors in the St. Jude Lifetime model-development cohort, including the clinical model variables and age at baseline, hypertension, genetic ancestry, and two general-population-derived polygenic risk scores

| Variables | Relative Risk<br>(95% CI) | P value |
| --- | --- | --- |
| Age at the primary diagnosis (years) |  |  |
| ≤5 | Reference |  |
| >5-10 | 0.96 (0.49-1.88) | 0.91 |
| >10-15 | 0.89 (0.46-1.72) | 0.73 |
| >15 | 0.77 (0.38-1.57) | 0.47 |
| Sex |  |  |
| Female | Reference |  |
| Male | 1.56 (0.98-2.49) | 0.061 |
| Cumulative Anthracycline dose (mg/m<sup>2</sup>)* |  |  |
| 0 | Reference |  |
| >0-100 | 0.98 (0.41-2.39) | 0.97 |
| >100-250 | 2.51 (1.31-4.82) | 0.0057 |
| >250 | 5.57 (3.04-10.21) | <0.001 |
| Mean heart radiation exposure dose (Gray) |  |  |
| None | Reference |  |
| >5 | 0.88 (0.48-1.61) | 0.69 |
| >5-15 | 1.41 (0.57-3.51) | 0.46 |
| >15-35 | 2.65 (1.38-5.09) | 0.0035 |
| >35 | 16.74 (6.24-44.90) | <0.001 |
| Age at baseline (years) |  |  |
| ≤25 | Reference |  |
| >25-35 | 1.19 (0.56-2.57) | 0.65 |
| >35-45 | 1.86 (0.84-4.12) | 0.12 |
| >45 | 2.49 (0.93-6.68) | 0.070 |
| Hypertension** |  |  |
| No | Reference |  |
| Yes | 2.26 (1.35-3.80) | 0.0017 |
| Genetic ancestry |  |  |
| European | Reference |  |
| African | 1.93 (0.98-3.80) | 0.060 |
| Other | 0.56 (0.08-4.14) | 0.57 |
| Polygenic risk scores |  |  |
| Hypertrophic cardiomyopathy | 0.88 (0.67-1.15) | 0.34 |
| Left ventricular end-systolic volume index | 1.15 (0.88-1.49) | 0.31 |

\*Doxorubicin toxicity equivalence
\*\*At baseline; CI, confidence interval

In SJLIFE model-development cohort, AUC of the clinical model 1 was 0.833 (95% CI=0.789-0.877) (Figure 1). Adding age at baseline (model 2), hypertension (model 3) and genetic ancestry (model 4) did not significantly improve the AUCs (0.844, *P*=0.15; 0.853, *P*=0.15; and 0.852, *P*=0.97, respectively). Using self-reported race instead of genetic ancestry yielded comparable results (Supplementary Table 5). Further adding two PRSs (PRS_LVESVi_ and PRS_HCM_) also did not significantly enhance the AUC (0.856, *P*=0.41) (Figure 1 and Supplementary Table 6). In CCSS model-validation cohort, the AUC of the clinical model 1 was 0.812 (95% CI=0.770-0.853). This AUC value remained unchanged with the additions of age at baseline (model 2), hypertension (model 3), and genetic ancestry (model 4) to model 1 (Figure 1). Similar results were observed when genetic ancestry was replaced by self-reported race (Supplementary Table 5). However, adding the two PRSs (model 5) significantly increased the AUC to 0.822 (95% CI=0.782-0.863; *P*=0.016) compared to model 4 (Figure 1 and Supplementary Table 6). Consistent results were observed among survivors of European ancestry in both SJLIFE model-development and CCSS model-validation cohorts (Supplementary Table 7). A sensitivity analysis showed that the AUC estimates for models 1-4 without genotype data in the entire CCSS cohort were comparable to those in CCSS-model validation cohort with genotype data (Supplementary Table 8).

**Figure 1.**
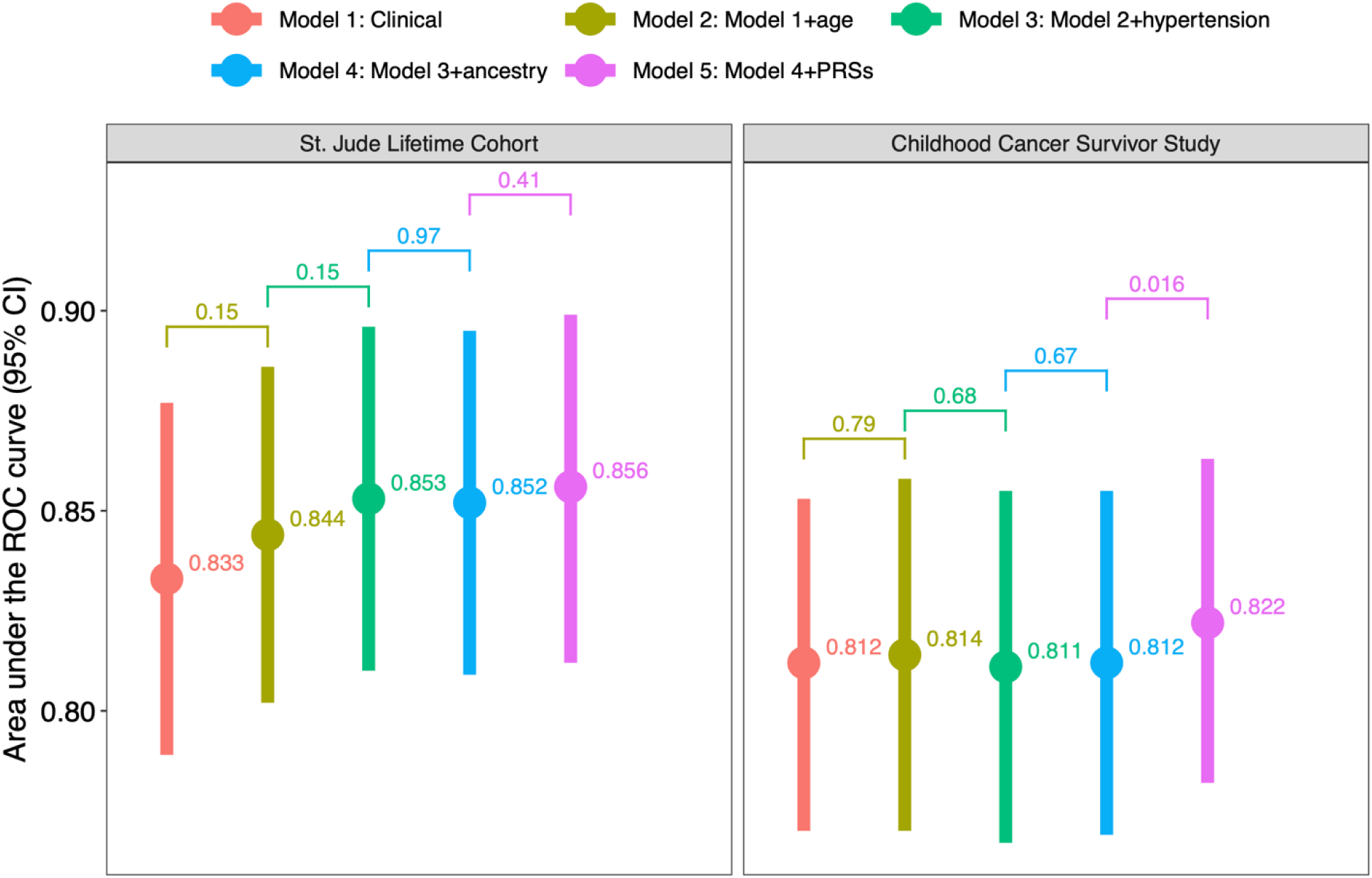
Performance of the risk prediction models for cardiomyopathy over the next 10 years from baseline in the St. Jude Lifetime model-development cohort and the Childhood Cancer Survivor Study model-validation cohort. Area under the ROC curve (AUC) estimates based the following five models – a baseline clinical model including demographic and treatment factors only; the baseline clinical model with age at baseline; the baseline clinical model with age at baseline and hypertension; the baseline clinical model with age at baseline, hypertension and genetic ancestry; and the baseline clinical model with age at baseline, CVRFs, genetic ancestry and two polygenic scores for hypertrophic cardiomyopathy and left ventricular end-systolic volume index developed from the general population.

In the subgroup of survivors exposed to cardiotoxic therapies, model 5 (which includes age at baseline, hypertension, genetic ancestry, and two PRSs) achieved an AUC of 0.844 (95% CI=0.796-0.892) in SJLIFE model-development cohort and 0.791 (95% CI=0.746-0.835) in CCSS model-validation cohort (Table 3). Adding these factors to the clinical model modestly improved prediction performance, though changes in AUC were not statistically significant. Similar results were observed in the IGHG-based moderate-risk group, with model 5 yielding an AUC of 0.836 (95% CI=0.754-0.917) in SJLIFE and 0.757 (95% CI=0.677-0.838) in CCSS. However, in the IGHG-based high and low-risk groups, the models did not further stratify risk among survivors.

**Table 3.** Performance of the cardiomyopathy risk prediction models among subpopulations of survivors exposed to cardiotoxic therapies and among those categorized into International Late Effects of Childhood Cancer Guideline Harmonization Group (IGHG)-based risk groups

| Survivor populations | Models | St Jude Lifetime Cohort |  | Childhood Cancer Survivor Study |  |
| --- | --- | --- | --- | --- | --- |
|  |  | AUC (95% CI) | P value | AUC (95% CI) | P value |
| Exposed to cardiotoxic therapies (Anthracyclines and/or heart radiation) | Model 1: Clinical | 0.810 (0.760-0.860) |  | 0.776 (0.732-0.820) |  |
|  | Model 2: Model 1 + age at baseline | 0.823 (0.775-0.871) | 0.14 | 0.784 (0.738-0.830) | 0.37 |
|  | Model 3: Model 2 + hypertension* | 0.835 (0.786-0.884) | 0.08 | 0.781 (0.735-0.827) | 0.70 |
|  | Model 4: Model 3 + genetic ancestry | 0.837 (0.788-0.885) | 0.76 | 0.781 (0.736-0.827) | 0.88 |
|  | Model 5: Model 4 + PRSs** | 0.844 (0.796-0.892) | 0.32 | 0.791 (0.746-0.835) | 0.39 |
| IGHG-based high-risk | Model 1: Clinical | 0.770 (0.693-0.847) |  | 0.538 (0.459-0.616) |  |
|  | Model 2: Model 1 + age at baseline | 0.741 (0.663-0.820) | 0.28 | 0.575 (0.492-0.659) | 0.074 |
|  | Model 3: Model 2 + hypertension* | 0.758 (0.679-0.837) | 0.32 | 0.585 (0.501-0.669) | 0.58 |
|  | Model 4: Model 3 + genetic ancestry | 0.758 (0.679-0.837) | 0.99 | 0.592 (0.509-0.674) | 0.44 |
|  | Model 5: Model 4 + PRSs** | 0.770 (0.696-0.843) | 0.57 | 0.628 (0.551-0.706) | 0.12 |
| IGHG-based moderate-risk | Model 1: Clinical | 0.729 (0.643-0.816) |  | 0.734 (0.633-0.835) |  |
|  | Model 2: Model 1 + age at baseline | 0.773 (0.689-0.857) | 0.027 | 0.753 (0.638-0.869) | 0.52 |
|  | Model 3: Model 2 + hypertension* | 0.808 (0.722-0.894) | 0.11 | 0.750 (0.665-0.834) | 0.91 |
|  | Model 4: Model 3 + genetic ancestry | 0.817 (0.731-0.904) | 0.46 | 0.747 (0.662-0.833) | 0.54 |
|  | Model 5: Model 4 + PRSs** | 0.836 (0.754-0.917) | 0.31 | 0.757 (0.677-0.838) | 0.76 |
| IGHG-based low-risk | Model 1: Clinical | 0.647 (0.500-0.795) |  | 0.476 (0.185-0.766) |  |
|  | Model 2: Model 1 + age at baseline | 0.689 (0.535-0.844) | 0.16 | 0.516 (0.224-0.808) | 0.71 |
|  | Model 3: Model 2 + hypertension* | 0.696 (0.533-0.860) | 0.79 | 0.471 (0.201-0.740) | 0.25 |
|  | Model 4: Model 3 + genetic ancestry | 0.701 (0.540-0.863) | 0.73 | 0.493 (0.235-0.752) | 0.084 |
|  | Model 5: Model 4 + PRSs** | 0.685 (0.527-0.844) | 0.34 | 0.510 (0.254-0.766) | 0.89 |

Clinical model (Model 1) included age at primary cancer diagnosis, sex, cumulative anthracycline dose and mean heart radiation exposure dose
\*At baseline
\*\*PRSs include those for hypertrophic cardiomyopathy and left ventricular end-systolic volume index developed from the general population
AUC, area under the receiver operating characteristic curve
CI, confidence interval
P values were obtained from the DeLong's test comparing AUC of adjacent models (for example, clinical + age at baseline vs. clinical)

Compared to the IGHG-based risk groups, the best-performing model (model 5) more effectively classified the 10-year risk of cardiomyopathy. It identified fewer survivors at high-risk (260 vs. 506) and moderate-risk (454 vs. 821) in SJLIFE, with similar trends in CCSS (high-risk: 361 vs. 1,476; moderate-risk: 856 vs. 1,566). At the same time, model 5 predicted more survivors at low-risk than IGHG guidelines. Notably, cumulative incidences of cardiomyopathy at 10 years from baseline were nearly 1.5 times higher in higher-risk groups based on model 5 compared to IGHG: specifically, in high-risk (SJLIFE: 23.9% vs. 16.7%; CCSS: 7.7% vs. 4.8%) and moderate-risk groups (SJLIFE: 9.9% vs. 6.2%; CCSS: 3.6% vs. 1.3%) (Figure 2). The low-risk group remained stable (SJLIFE: 2.3% vs. 2.0%; CCSS: 1.0% vs. 0.4%). While model 5 showed good risk discrimination, the CCSS dataset indicated slight overestimation in risk: high and moderate-risk groups were calibrated to be >10% and 5%≤predicted probability≤10%, but CCSS showed 7.7% and 3.6%, respectively.

**Figure 2.**
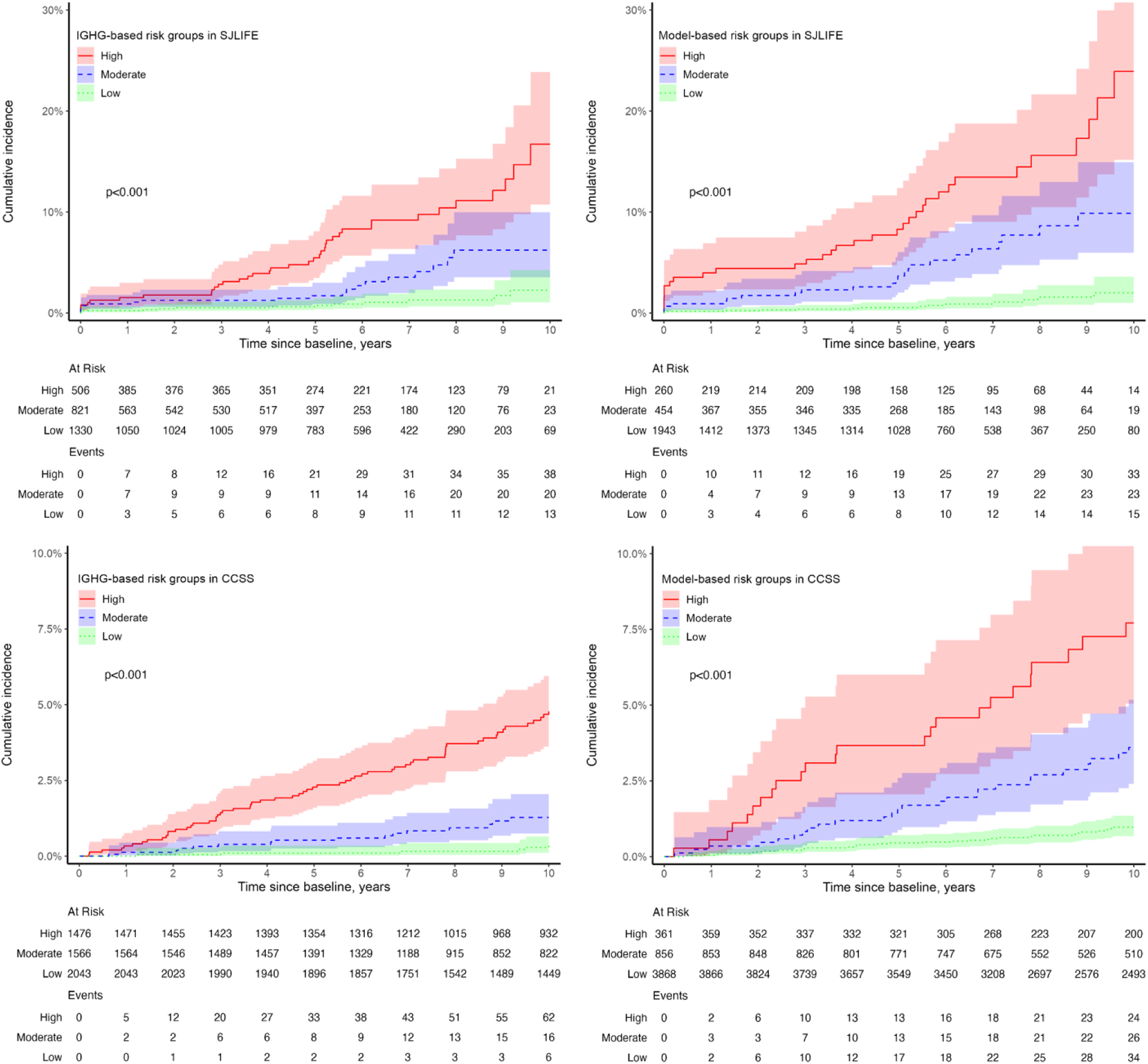
Cumulative incidences of cardiomyopathy over the next 10 years from baseline in the St. Jude Lifetime Cohort (SJLIFE) and Childhood Cancer Survivor Study (CCSS). Estimates are shown by different risk groups defined by the International Guideline Harmonization Group (panels A [SJLIFE] and C [CCSS]) and based on predicted counts from the best-performing model that included demographic and cancer treatment exposures, age at baseline, hypertension, genetic ancestry and the two polygenic risk scores for hypertrophic cardiomyopathy and left ventricular end-systolic volume index from the general population (panels B [SJLIFE] and D [CCSS]). P values were calculated using Gray’s test comparing cumulative incidences among the three risk groups. The y-axis scales were adjusted between SJLIFE and CCSS cohorts to accommodate differences in cumulative incidences and improve visual clarity.

## DISCUSSION

Using a large, clinically-assessed cohort of long-term survivors of childhood cancer, we developed a prediction model for estimating the 10-year risk of cardiomyopathy. The model, including cancer treatments, age, hypertension and two PRSs developed from the general population, demonstrated good discrimination in both model-development (AUC 0.856) and model-validation cohorts (AUC 0.822), highlighting its clinical utility. Notably, this model was more effective in identifying high- and moderate-risk survivors compared to IGHG guidelines. It is available as an online risk calculator at https://ccss.stjude.org/resources/calculators/cardiomyopathy-prediction-calculator.html.

Our risk prediction model, based on clinically-assessed outcomes, showed higher AUC values than those using self-reported data among CCSS survivors^14,15^. While our model targeted the 10-year risk from baseline assessment (with most survivors being adults at baseline), earlier CCSS models focused on risk after 5 years post-childhood cancer diagnosis. Chow et al.^14^ developed a model for predicting HF after 5-year survival before age 40, using data from 13,060 survivors, with an AUC of 0.76. Chen et al.^15^ expanded this by including traditional CVRFs and predicting HF from ages 20, 25, 30, and 35 by age 50, yielding AUCs between 0.69 and 0.77. Unlike these previous models, which were set-up to predict HF by a certain age and therefore, did not include attained age as a predictor, our model was established for use in current long-term survivors who are substantially older at the time of prediction and the models were designed for predicting cardiomyopathy over the next 10 years. Thus, age at the time of prediction was included to account for age-related risk.

The AUC of our clinical model 1, with demographic and treatment factors, increased by 2% in the model-development cohort when age, hypertension, and genetic ancestry were added, but showed no change in the model-validation cohort. Age and hypertension were associated with higher cardiomyopathy rates in the model-development cohort, however, and genetic ancestry showed marginal evidence of association. Consistent with the observed associations, the cardiovascular disease risk and incidence of CVRFs are known to increase with survivors’ age^40^. Furthermore, age is a well-established risk factor for cardiovascular disease and is included in most cardiovascular risk scores of the general population^41,42^. Survivors of African genetic ancestry or self-reported Black race are known to have significantly higher cardiomyopathy rates compared to European genetic ancestry or self-reported White race counterparts,^21^ which underscores the importance of including this factor in risk prediction models.

Two PRSs from the general population enhanced the 10-year cardiomyopathy risk prediction in the model-validation cohort, even though neither showed statistically significant association with cardiomyopathy rate in the model-development cohort. The PRS for hypertrophic cardiomyopathy was associated with lower risk of cardiomyopathy in our model-development cohort, consistent with the UK Biobank findings that hypertrophic and dilated cardiomyopathy share genetic pathways with opposite effect directions^16^. The observed association of the PRS for left ventricular end-systolic volume was consistent with that of the UK Biobank where the risks of dilated cardiomyopathy^18^ and HF^43^ were found to be associated with this PRS. This may suggest shared genetic mechanisms for certain cardiac changes in childhood cancer survivors and the general population.

Currently, IGHG estimates cardiomyopathy risk in childhood cancer survivors based solely on cardiotoxic treatment exposures. In contrast, our best-performing model 5, which includes two PRSs, more effectively predicted the 10-year risk of cardiomyopathy, specifically in high and moderate-risk groups. Compared to IGHG, it classified fewer survivors as high-risk and moderate-risk and identified more as low-risk. Notably, the cumulative incidence of cardiomyopathy was 1.5-times higher in the high-risk and moderate-risk groups identified by our model than by IGHG. Both IGHG and the Children’s Oncology Group recommend routine echocardiography every two years for high-risk and every five years for moderate-risk survivors, while it is no longer recommended for low-risk survivors ^38^. We defined low-risk survivors as those with a <5% predicted probability of developing cardiomyopathy over the next 10 years, observing 2.3% in SJLIFE and 1.0% in CCSS in this group. Leerink and colleagues have proposed a similar re-stratification approach using real-time risk estimates from echocardiogram results, suggesting potential reductions in screening intensity for some survivors^44^. If validated by cost-effectiveness analyses, our model could enhance risk stratification for cardiomyopathy in adult survivors of childhood cancer and potentially lower screening costs for high- and moderate-risk individuals.

Our study has several limitations. First, we did not consider lifestyle factors such as smoking, alcohol consumption, obesity, physical activity, and diet. However, the inclusion of CVRFs, which are often more common in individuals with suboptimal lifestyles, may reflect some of these behaviors. Second, some PRSs were based on a limited number of variants, potentially missing the full extent of genetic effects. Third, our model was not well calibrated in the CCSS validation cohort (Supplementary Figure 1), where the cardiomyopathy rate was consistently lower than in SJLIFE, with a multiplicative factor of 0.6, despite using the same risk thresholds. This uniformly lower risk suggests under-ascertainment or under-reporting of cardiomyopathy in CCSS and does not necessarily suggest poor prediction or lower risk in CCSS survivors. Fourth, while our model adjusted for genetic ancestry, over 80% of survivors were of European ancestry, which may limit its generalizability to non-European ancestry survivors. Given the higher risk of cardiomyopathy in African American survivors^21^, it is essential to collect data and develop race-specific risk prediction models for minority groups.

In conclusion, we developed and validated a prediction model to estimate the 10-year risk of cardiomyopathy in childhood cancer survivors. This model incorporates cancer treatments, age, hypertension, ancestry, and PRSs from the general population, resulting in improved accuracy in risk classification. It more effectively identified at-risk survivors compared to the current IGHG-based groups recommended for cardiac surveillance. With the highest prediction performance to date based on clinically assessed outcomes, our model is well-suited for long-term survivors facing age-related risks and can help identify those who would benefit most from preventive interventions and risk reduction strategies.

## Data Availability

All data produced in the present study are available upon reasonable request to the authors

https://ccss.stjude.org/resources/calculators/cardiomyopathy-prediction-calculator.html

