## Supplemental material for "Predicting the 10-year risk of cardiomyopathy in long-term survivors of childhood cancer: a report from the St. Jude Lifetime Cohort and the Childhood Cancer Survivor Study"

**SUPPLEMENTARY METHODS**

**Cancer therapy exposures**

Information pertaining to exposures to chemotherapy and radiotherapy was abstracted from medical records. The cumulative anthracycline dose (expressed in mg/m^2^) was determined by doxorubicin toxicity equivalence^1^. A centralized review was conducted for the radiotherapy records by radiation physicists at the MD Anderson Cancer Center in Houston, TX, and the mean radiation dose to the heart for each survivor was calculated using established radiation dosimetry methodologies and included reconstructing their radiotherapy fields on a computational phantom, scaled to the age at treatment^2-4^.

**Genotype data**

Methods used to generate genotype data in SJLIFE and CCSS have been extensively described previously^5-7^. Briefly, whole genomes of SJLIFE survivors and a subset of CCSS survivors diagnosed between 1987 and 1999 were sequenced using the Illumina HiSeq X10 and/or NovaSeq platforms with 30X average coverage per sample^8,9^. Genotype data of CCSS survivors diagnosed between 1970 and 1986 were obtained on Illumina HumanOmni5Exome array and the genotypes were imputed up to the Haplotype Reference Consortium r1.1 (2016) haplotypes using Minimac3^10^, implemented in the Michigan Imputation Server^10,11^. Prior to the analyses, stringent sample and variant quality control procedures were applied to the genotype data in both cohorts, as described earlier^5,12-14^. For each survivor in SJLIFE and CCSS, European, African, and East Asian genetic ancestry was estimated using ADMIXTURE^15^ on the basis of genotype data of an independent set of common autosomal variants and the 1000 Genomes Project samples as ancestral populations. Survivors were grouped into European (%European >80%)^16^, African (%African >60%)^17^ and Other based on the estimated ancestry proportions.

**General population polygenic risk scores**

Based on the published GWASs in the general population, we considered multiple PRSs for cardiomyopathies (dilated [DCM] and hypertrophic [HCM]), heart failure (HF), measures of cardiac structure and function that are predictive of cardiac dysfunction, and anthracycline-induced cardiotoxicity. The first PRS included 13 variants associated with the risk of dilated cardiomyopathy in a recent GWAS among 19,260 White participants from the UK Biobank^18^. Results from the same study also found a significant association of a 20-variant PRS for hypertrophic cardiomyopathy (PGS catalog: PGS000778) with LV wall thickness and adverse cardiac events in an independent sample. We assessed this PRS considering strong genetic correlations between dilated and hypertrophic cardiomyopathies and left ventricular traits^18^. We also considered a PRS including 910,146 variants for HF (PGS catalog: PGS001790) constructed and validated among individuals of diverse ancesries^19^. In another GWAS for cardiac magnetic resonance imaging-derived LV traits among 36,000 White UK Biobank participants, PRSs for LVEF (22 variants) and LV end systolic volume index (LVESVi; 28 variants) were strongly associated with incident dilated cardiomyopathy: we considered both PRSs^20^. Finally, we also constructed a PRS based on seven SNPs previously associated with the risk of anthracycline-induced cardiotoxicity^12,13,21-24^. Using the reported weights of these variants (Supplementary Table 2), six separate PRSs (PRS_DCM_, PRS_HCM_, PRS_HF,_ PRS_LVEF_, PRS_LVESVi_, and PRS_ACT_) were constructed as weighted sums of the number of risk alleles carried by each survivor.

Supplementary Table 1 | Common Terminology Criteria for Adverse Events (CTCAE) grading of cardiomyopathy (St. Jude Lifetime) and heart failure (Childhood Cancer Survivor Study).

| **CTCAE grades** | **St. Jude Lifetime Cohort** | **Childhood Cancer Survivor Study** |
| --- | --- | --- |
| 2 (Moderate) | Resting EF < 50-40%; 10 - 19% absolute drop from baseline | Congestive heart failure, not requiring medication |
| 3 (Severe) | Resting EF 39-20%; >20% absolute drop from baseline; medication indicated or initiated | Cardiomyopathy or congestive heart failure requiring medication |
| 4 (Life-threatening) | Resting EF<20%; refractory or poorly controlled heart failure due to drop in ejection fraction; on medical management; intervention such as ventricular assist device, intravenous vasopressor support, or heart transplant indicated | Cardiac transplantation |
| 5 (Death) | Death | Death as a result of heart failure |

Supplementary Table 2 | Genetic variants identified in the general population used for constructing polygenic risk scores in childhood cancer survivors from the St. Jude Lifetime model-development cohort and the Childhood Cancer Survivor Study model-validation cohort

| **Chromosome** | **Position (hg38)** | **Effect allele** | **Non-effect allele** | **log-odds/beta estimate** |
| --- | --- | --- | --- | --- |
| *Dilated cardiomyopathy* | | | | |
| 1 | 15972817 | C | T | 0.13 |
| 7 | 128848680 | G | A | 0.15 |
| 10 | 119670121 | T | C | 0.19 |
| 2 | 178693639 | T | C | 0.11 |
| 3 | 14232951 | G | A | 0.08 |
| 6 | 36665292 | G | A | 0.10 |
| 8 | 11922131 | G | A | 0.09 |
| 8 | 124845117 | G | T | 0.10 |
| 10 | 29418622 | G | A | 0.09 |
| 15 | 84657289 | T | G | 0.09 |
| 17 | 66307675 | C | T | 0.08 |
| 18 | 36744128 | G | A | 0.08 |
| 22 | 23829118 | G | A | 0.11 |
| *Hypertrophic cardiomyopathy* | | | | |
| 1 | 16032437 | T | C | 0.10 |
| 2 | 36923650 | G | A | 0.07 |
| 3 | 69981390 | T | A | 0.08 |
| 3 | 172067378 | C | G | 0.07 |
| 5 | 57715642 | T | A | 0.09 |
| 5 | 139407567 | A | G | 0.08 |
| 6 | 36668303 | C | T | 0.13 |
| 6 | 118331912 | A | G | 0.17 |
| 7 | 128790383 | T | G | 0.13 |
| 10 | 73646383 | C | A | 0.12 |
| 10 | 112728053 | A | G | 0.09 |
| 10 | 119659975 | G | C | 0.14 |
| 11 | 14045253 | T | G | 0.07 |
| 15 | 84806850 | C | T | 0.10 |
| 16 | 927160 | C | T | 0.08 |
| 17 | 1392798 | G | C | 0.12 |
| 17 | 46742199 | T | C | 0.11 |
| 17 | 66307675 | T | C | 0.08 |
| 18 | 36673782 | C | T | 0.13 |
| 22 | 23829118 | A | G | 0.12 |
| *Left ventricular end systolic volume (body surface area-indexed)* | | | | |
| 1 | 6212077 | G | A | 0.04 |
| 1 | 16021917 | C | T | 0.07 |
| 1 | 45541360 | C | CAA | 0.04 |
| 2 | 178649706 | T | C | 0.09 |
| 2 | 179229625 | C | T | 0.37 |
| 2 | 200333900 | C | CATT | 0.04 |
| 3 | 14231266 | G | C | 0.06 |
| 3 | 69807602 | C | T | 0.05 |
| 3 | 158588625 | A | G | 0.04 |
| 3 | 172042637 | T | C | 0.04 |
| 5 | 139394348 | T | C | 0.04 |
| 6 | 36679512 | G | A | 0.07 |
| 7 | 128846309 | C | T | 0.07 |
| 8 | 124846296 | GA | G | 0.05 |
| 8 | 140625230 | C | T | 0.04 |
| 10 | 110784367 | C | T | 0.21 |
| 10 | 119656173 | G | A | 0.09 |
| 11 | 14042845 | A | G | 0.04 |
| 12 | 120230731 | A | G | 0.13 |
| 15 | 84805730 | T | TTTTG | 0.05 |
| 16 | 942961 | T | G | 0.05 |
| 17 | 1470901 | G | A | 0.07 |
| 17 | 45949373 | A | G | 0.04 |
| 17 | 55297249 | T | C | 0.06 |
| 17 | 66307675 | C | T | 0.04 |
| 18 | 58289633 | G | T | 0.04 |
| 19 | 45812551 | G | A | 0.05 |
| 22 | 23836092 | A | G | 0.06 |
| *Left ventricular ejection fraction* | | | | |
| 1 | 2212668 | G | A | 0.06 |
| 1 | 16011438 | C | T | 0.07 |
| 1 | 236678777 | C | G | 0.04 |
| 2 | 178649706 | C | T | 0.08 |
| 3 | 14250179 | C | T | 0.07 |
| 3 | 69808622 | T | C | 0.06 |
| 5 | 139421136 | A | G | 0.05 |
| 6 | 32668263 | C | T | 0.04 |
| 6 | 36679512 | A | G | 0.08 |
| 7 | 128832084 | A | G | 0.08 |
| 8 | 11929416 | T | C | 0.04 |
| 8 | 140694133 | T | C | 0.04 |
| 10 | 110784367 | T | C | 0.20 |
| 10 | 119656173 | A | G | 0.10 |
| 11 | 19191179 | T | A | 0.07 |
| 12 | 26195496 | C | G | 0.05 |
| 15 | 84779989 | T | G | 0.05 |
| 16 | 938070 | A | T | 0.04 |
| 17 | 55297249 | C | T | 0.06 |
| 18 | 36604896 | C | T | 0.04 |
| 18 | 58289633 | T | G | 0.04 |
| 22 | 23817120 | A | T | 0.06 |
| *Anthracycline-related cardiomyopathy* | | | | |
| 1 | 114101719.00 | C | T | 1.69 |
| 2 | 234602277.00 | A | C | 1.46 |
| 6 | 39262535.00 | A | C | 0.59 |
| 6 | 43272188.00 | A | G | 0.80 |
| 9 | 86900926.00 | G | A | 1.02 |
| 12 | 53605545.00 | A | G | 1.55 |
| 14 | 23814569.00 | G | A | 0.69 |

Supplementary Table 3 | Multivariable Poisson regression models estimating the cardiomyopathy incidence rate over the next 10 years from baseline among childhood cancer survivors in the St. Jude Lifetime model-development cohort, including the clinical model variables and age at baseline, hypertension and genetic ancestry

| **Variables** | **Model 1**  **(Clinical)** | | **Model 2**  **(Model 1+age at baseline)** | | **Model 3**  **(Model 2+hypertension** | | **Model 4**  **(Model 3+genetic ancestry)** | |
| --- | --- | --- | --- | --- | --- | --- | --- | --- |
|  | **RR (95% CI)** | **P value** | **RR (95% CI)** | **P value** | **RR (95% CI)** | **P value** | **RR (95% CI)** | **P value** |
| Age at primary diagnosis (years) |  |  |  |  |  |  |  |  |
| ≤5 | Reference | | | | | | | |
| >5-10 | 1.06 (0.55 - 2.06) | 0.86 | 0.97 (0.50 - 1.90) | 0.94 | 0.97 (0.50 - 1.89) | 0.93 | 0.98 (0.50 - 1.91) | 0.95 |
| >10-15 | 1.16 (0.62 - 2.15) | 0.65 | 0.88 (0.45 - 1.70) | 0.71 | 0.91 (0.47 - 1.75) | 0.77 | 0.90 (0.46 - 1.73) | 0.74 |
| >15 | 1.21 (0.63 - 2.32) | 0.57 | 0.81 (0.40 - 1.65) | 0.56 | 0.79 (0.39 - 1.61) | 0.51 | 0.79 (0.39 - 1.61) | 0.51 |
| Sex |  |  |  |  |  |  |  |  |
| Female | Reference | | | | | | | |
| Male | 1.53 (0.96 - 2.44) | 0.072 | 1.55 (0.97 - 2.48) | 0.064 | 1.54 (0.97 - 2.47) | 0.069 | 1.54 (0.96 - 2.45) | 0.070 |
| Cumulative anthracycline dose (mg/m^2^)* |  |  |  |  |  |  |  |  |
| None | Reference | | | | | | | |
| >0-100 | 0.77 (0.32 - 1.82) | 0.55 | 0.93 (0.39 - 2.25) | 0.87 | 0.94 (0.39 - 2.26) | 0.88 | 0.98 (0.40 - 2.36) | 0.96 |
| >100-250 | 1.79 (0.97 - 3.31) | 0.063 | 2.30 (1.20 - 4.40) | 0.012 | 2.36 (1.24 - 4.51) | 0.0092 | 2.48 (1.29 - 4.75) | 0.0064 |
| >250 | 4.23 (2.39 - 7.50) | <.001 | 4.86 (2.68 - 8.80) | <.001 | 5.15 (2.84 - 9.35) | <.001 | 5.41 (2.97 - 9.87) | <.001 |
| Mean heart radiation exposure dose (Gray) |  |  |  |  |  |  |  |  |
| None | Reference | | | | | | | |
| >0-5 | 1.09 (0.61 - 1.96) | 0.78 | 0.92 (0.50 - 1.68) | 0.79 | 0.89 (0.49 - 1.63) | 0.71 | 0.89 (0.49 - 1.62) | 0.70 |
| >5-15 | 1.57 (0.64 - 3.87) | 0.33 | 1.44 (0.58 - 3.59) | 0.43 | 1.42 (0.57 - 3.54) | 0.45 | 1.42 (0.57 - 3.54) | 0.45 |
| >15-35 | 2.62 (1.37 - 5.01) | 0.0037 | 2.49 (1.30 - 4.77) | 0.0060 | 2.65 (1.38 - 5.10) | 0.0034 | 2.62 (1.36 - 5.03) | 0.0038 |
| >35 | 17.84 (7.08 - 44.97) | <.001 | 14.75 (5.54 - 39.27) | <.001 | 16.66 (6.26 - 44.34) | <.001 | 15.80 (5.91 - 42.20) | <.001 |
| Age at baseline |  |  |  |  |  |  |  |  |
| ≤25 | Reference | | | | | | | |
| >25-35 |  |  | 1.33 (0.62 - 2.86) | 0.46 | 1.24 (0.58 - 2.68) | 0.58 | 1.24 (0.58 - 2.66) | 0.59 |
| >35-45 |  |  | 2.40 (1.10 - 5.20) | 0.027 | 1.96 (0.88 - 4.34) | 0.10 | 1.96 (0.89 - 4.33) | 0.095 |
| >45 |  |  | 3.35 (1.27 - 8.78) | 0.014 | 2.47 (0.92 - 6.63) | 0.073 | 2.47 (0.92 - 6.62) | 0.073 |
| Hypertension |  |  |  |  |  |  |  |  |
| No | Reference | | | | | | | |
| Yes |  |  |  |  | 2.20 (1.31 - 3.67) | 0.0027 | 2.19 (1.31 - 3.65) | 0.0027 |
| Genetic ancestry |  |  |  |  |  |  |  |  |
| European | Reference | | | | | | | |
| African |  |  |  |  |  |  | 1.70 (0.94 - 3.06) | 0.078 |
| Others |  |  |  |  |  |  | 0.54 (0.07 - 3.92) | 0.54 |

*Doxorubicin toxicity equivalence

CI, confidence interval

Clinical model (Model 1) included age at primary cancer diagnosis, sex, cumulative anthracycline dose and mean heart radiation dose

Supplementary Table 4 | Associations of the general population polygenic risk scores with the rate of cardiomyopathy over the next 10 years from baseline among survivors in the St. Jude Lifetime Cohort model-development cohort

| **Polygenic risk scores** | **Relative Rate**  **(95% CI)** | **P value** |
| --- | --- | --- |
| *All survivors* | | |
| Dilated Cardiomyopathy | 1.01 (0.80-1.28) | 0.92 |
| Hypertrophic cardiomyopathy | 0.82 (0.65-1.05) | 0.12 |
| Left ventricular ejection fraction | 0.98 (0.76-1.26) | 0.88 |
| Left ventricular end-systolic volume index | 1.21 (0.95-1.54) | 0.11 |
| Anthracycline-related cardiotoxicity | 1.19 (0.95-1.49) | 0.13 |
| Heart failure | 0.95 (0.74-1.22) | 0.68 |
| *Survivors of European ancestry* | | |
| Dilated Cardiomyopathy | 1.07 (0.83-1.37) | 0.62 |
| Hypertrophic cardiomyopathy | 0.72 (0.54-0.96) | 0.024 |
| Left ventricular ejection fraction | 0.97 (0.73-1.27) | 0.81 |
| Left ventricular end-systolic volume index | 1.26 (0.97-1.64) | 0.082 |
| Anthracycline-related cardiotoxicity | 1.11 (0.86-1.44) | 0.41 |
| Heart failure | 0.92 (0.70-1.21) | 0.56 |

Results were adjusted for age at primary cancer diagnosis, sex, cumulative anthracycline dose, mean heart radiation exposure dose, age at baseline, hypertension and genetic ancestry (only for analysis including all survivors)

Supplementary Table 5 | Performance of the cardiomyopathy risk prediction models estimating the cardiomyopathy incidence rate over the next 10 years from baseline among survivors in the St. Jude Lifetime model-development cohort and Childhood Cancer Survivor Study model-validation cohort, replacing genetic ancestry by self-reported race

| **Models** | **St. Jude**  **Lifetime Cohort**  **(Discovery Cohort)** | | **Childhood Cancer**  **Survivor Study**  **(Validation Cohort)** | |
| --- | --- | --- | --- | --- |
|  | **AUC (95% CI)** | **P value** | **AUC (95% CI)** | **P value** |
| Model 1: Clinical | 0.833 (0.789-0.877) |  | 0.812 (0.770-0.853) |  |
| Model 2: Clinical + age at baseline | 0.844 (0.802-0.886) | 0.15 | 0.814 (0.770-0.858) | 0.79 |
| Model 3: Clinical + age at baseline + hypertension | 0.853 (0.810-0.896) | 0.15 | 0.811 (0.767-0.855) | 0.68 |
| Model 4: Clinical + age at baseline + hypertension + race | 0.853 (0.811-0.886) | 0.85 | 0.810 (0.764-0.855) | 0.53 |

Clinical model included age at primary cancer diagnosis, sex, cumulative anthracycline dose and mean heart radiation exposure dose

AUC, area under the receiver operating characteristic curve

CI, confidence interval

P values were obtained from the DeLong’s test comparing AUC of adjacent models (for example, clinical + age at baseline vs. clinical)

Supplementary Table 6 | Performance of the cardiomyopathy risk prediction models estimating the cardiomyopathy incidence rate over the next 10 years from baseline among all survivors in the St. Jude Lifetime model-development cohort and Childhood Cancer Survivor Study model-validation cohort, including polygenic risk scores from the general population

| **Models** | **St. Jude Lifetime Cohort** | |  | **Childhood Cancer Survivor Study** | |
| --- | --- | --- | --- | --- | --- |
|  | **AUC (95% CI)** | **P value** |  | **AUC (95% CI)** | **P value** |
| Model 4: Clinical + age at baseline + hypertension + genetic ancestry | 0.852 (0.809-0.895) |  |  | 0.812 (0.769-0.855) |  |
| Model 4 + PRS_HCM_ | 0.854 (0.810-0.898) | 0.68 |  | 0.820 (0.777-0.862) | 0.085 |
| Model 4 + PRS_LVESVi_ | 0.856 (0.813-0.899) | 0.31 |  | 0.822 (0.782-0.862) | 0.026 |
| Model 5: Model 4 + PRS_HCM_ + PRS_LVESVi_ | 0.856 (0.812-0.899) | 0.41 |  | 0.824 (0.784-0.865) | 0.016 |

Clinical model included age at primary cancer diagnosis, sex, cumulative anthracycline dose and mean heart radiation exposure dose

PRS_HCM_, polygenic risk score for hypertrophic cardiomyopathy

PRS_LVESVi_, polygenic risk score for left ventricular end systolic volume index

AUC, area under the receiver operating characteristic curve

CI, confidence interval

P values were obtained from the DeLong’s test comparing AUC of models with one or more PRSs with the AUC of clinical model with attained age, hypertension and genetic ancestry

Supplementary Table 7 | Performance of the cardiomyopathy risk prediction models estimating the cardiomyopathy incidence rate over the next 10 years from baseline among survivors of European ancestry in the St. Jude Lifetime model-development cohort and Childhood Cancer Survivor Study model-validation cohort, including polygenic risk scores from the general population

| **Models** | **St. Jude Lifetime Cohort** | |  | **Childhood Cancer Survivor Study** | |
| --- | --- | --- | --- | --- | --- |
|  | **AUC (95% CI)** | **P value** |  | **AUC (95% CI)** | **P value** |
| Clinical + age at baseline + hypertension | 0.838 (0.785-0.890) |  |  | 0.794 (0.750-0.839) |  |
| Clinical + age at baseline + hypertension + PRS_HCM_ | 0.842 (0.789-0.894) | 0.57 |  | 0.798 (0.753-0.843) | 0.66 |
| Clinical + age at baseline + hypertension + PRS_LVESVi_ | 0.841 (0.788-0.893) | 0.58 |  | 0.804 (0.762-0.846) | 0.11 |
| Clinical + age at baseline + hypertension + PRS_HCM_ + PRS_LVESVi_ | 0.842 (0.790-0.895) | 0.53 |  | 0.803 (0.760-0.846) | 0.30 |

Clinical model included age at primary cancer diagnosis, sex, cumulative anthracycline dose and mean heart radiation exposure dose

PRS_HCM_, polygenic risk score for hypertrophic cardiomyopathy

PRS_LVESVi_, polygenic risk score for left ventricular end systolic volume index

AUC, area under the receiver operating characteristic curve

CI, confidence interval

P values were obtained from the DeLong’s test comparing AUC of models with one or more PRSs with the AUC of clinical model with attained age and hypertension

Supplementary Table 8 | Performance of the cardiomyopathy risk prediction models without requiring genotype data estimating the cardiomyopathy incidence rate over the next 10 years from baseline developed among survivors in the St. Jude Lifetime model-development cohort, in the entire Childhood Cancer Survivor Study

| **Models** | **AUC (95% CI)** | **P value** |
| --- | --- | --- |
| Model 1: Clinical | 0.792 (0.759-0.824) |  |
| Model 2: Clinical + age at baseline | 0.792 (0.758-0.825) | 0.99 |
| Model 3: Clinical + age at baseline + hypertension | 0.793 (0.760-0.826) | 0.74 |
| Model 4: Clinical + age at baseline + hypertension + race | 0.785 (0.751-0.819) | <.001 |

Clinical model included age at primary cancer diagnosis, sex, cumulative anthracycline dose and mean heart radiation exposure dose

AUC, area under the receiver operating characteristic curve

CI, confidence interval

P values were obtained from the DeLong’s test comparing AUC of adjacent models (for example, clinical + age at baseline vs. clinical)

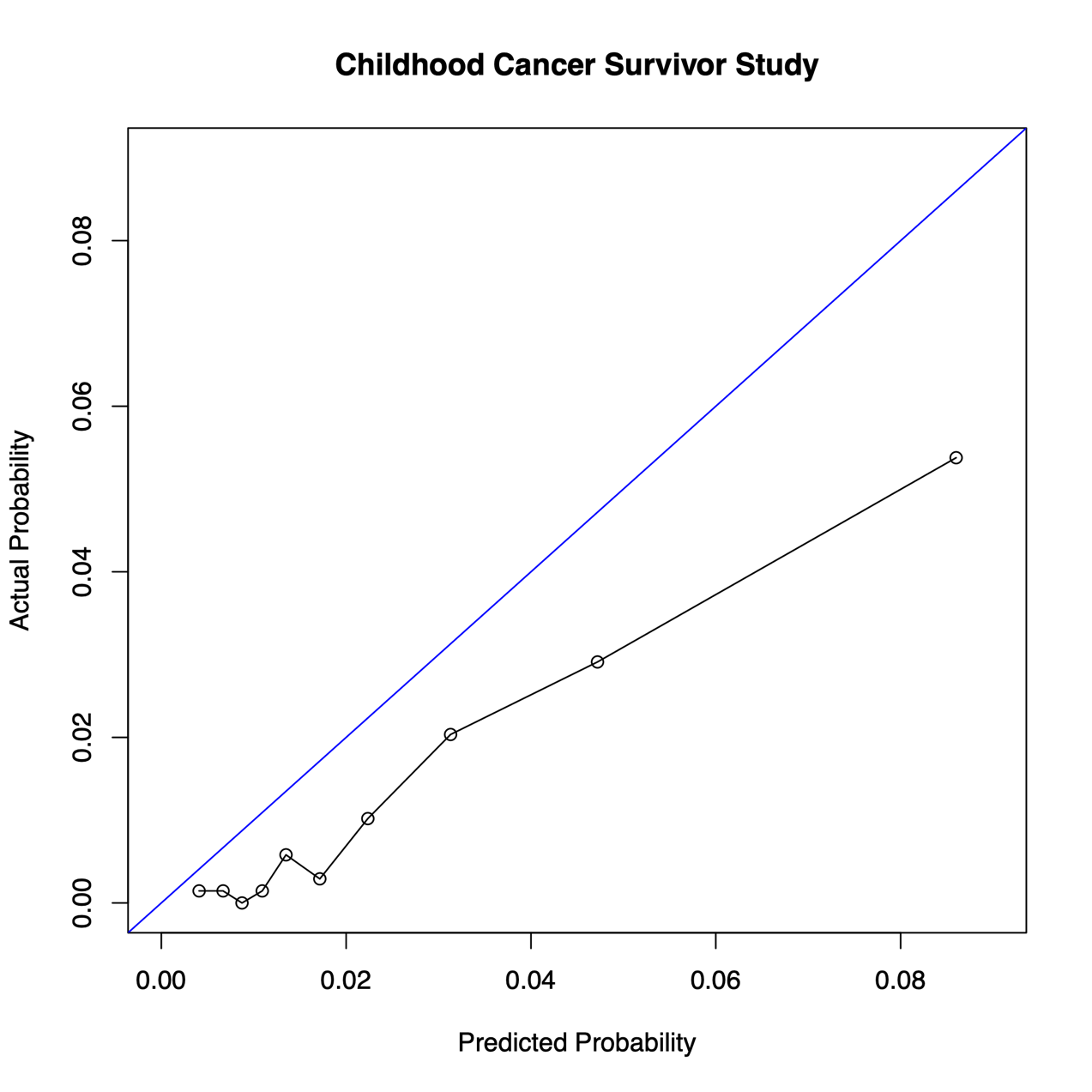

Supplementary Figure 1 | Calibration plot showing actual vs. predicted probability of developing cardiomyopathy over the next 10 years from baseline in the Childhood Cancer Survivor Study model-validation cohort, based on the best-performing model 5 including demographic, cancer treatment exposures, age at baseline, hypertension, genetic ancestry and the two polygenic scores for hypertrophic cardiomyopathy and left ventricular end-systolic volume index from the general population.
